# COVID-19 in Canada: Predictions for the future and control lessons from Asia

**DOI:** 10.1101/2020.03.21.20040667

**Authors:** Cornelius Christian, Francis Christian

## Abstract

COVID-19 has spread with unequal efficiency in various parts of the world. In several European countries including Italy, the increase in the number of COVID-19 cases has followed a consistent, exponential pattern of spread. However, some countries, notably Taiwan and Hong Kong, have achieved a different outcome and have managed to bring the COVID-19 outbreak in their countries rapidly under control, without entering the exponential pattern and with very few cases. They have used several different approaches to COVID-19 outbreak control, including the innovative use of smartphone technology and the widespread use of surgical face masks. We show through our models, that Canada has followed the same, consistent COVID-19 exponential growth pattern that is seen in Italy. Both nationally and in its most heavily affected provinces, there is exponential growth of COVID-19 cases, making it possible to make predictions for the future, if no further interventions are made in public health policy. In particular, we argue for the urgent introduction of surgical face masks in health care and other settings and the harnessing of the power of smartphone technology on a national scale.

---

In December 2019, a novel coronavirus started in a small corner of Wuhan, China, and spread rapidly across the globe. There are now more than 270,000 confirmed cases of COVID-19 worldwide, with over 11,000 deaths.^1^ On March 11, 2020, the World Health Organization declared the virus a pandemic, suggesting the need for an international and coordinated response. Such a response requires a proactive attitude, with policymakers making decisions based on real-time data and mathematical modelling.

Canada, like most countries, was caught unawares by the virus, causing key decision-makers to rush to action: school closures, bans on public gatherings, and travel restrictions are among the non-pharmaceutical interventions (NPIs) that have been adopted.^2, 3^

Like many other European and North American countries, Canada is constrained by limited ICU beds, which could curtail its ability to contain the virus; in normal times, the country’s large and teaching hospitals’ ICUs operate at 90×0025; capacity, and demand for ventilation units has grown over the past decade.^4^ These factors can potentially impede Canada’s ability to manage COVID-19.

In order to help assist policymakers, we present an exponential fit of Canadian COVID-19 cases, as a replication of,^5^ in order to provide a prediction of what could happen to coronavirus case numbers. Given what we know about European and Asian cases, we integrate these countries into our discussion in order to suggest appropriate NPIs for Canadian federal and provincial policymakers.

Based on these models and predictions, we also question the reluctance of Canadian policy makers to learn from jurisdictions such as Taiwan and Hong Kong, where the COVID-19 outbreak appears to have been brought under control.^6^ These countries have also extensively adopted face masks for the wider public, which along with hand hygiene are demonstrated to be effective against viral spread.^7, 8^

## Methodology and Results

In order to predict the trajectory of COVID-19 in Canada, we fit an exponential model to data on COVID-19 cases, in line with prior literature, which suggests that an epidemic’s early stages can be exponential.^9^ This fit is conducted via non-linear least squares estimation using the statistical package R. In particular, consider the following exponential model:

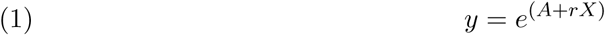

where *y* is the number of confirmed COVID-19 cases, and *X* is the number of days that the region has had the novel coronavirus. *A* and *r* are parameter values that we use non-linear least squares regression to fit.

The number of those infected with COVID-19 has been published daily since January 25, 2020. Based on these data from government sources, we can calculate the exponential fit for these cases, as seen in Appendix, Figure 1. The parameter values from this fit are shown in Table 1.

**Table 1.** Parameter Estimates of Exponential Model

| Parameter | Estimate | 95% CI |
| --- | --- | --- |
| $A_{CAN}$ | -6.247 | [-6.502, -5.991] |
| $r_{CAN}$ | 0.237 | [0.232, 0.241] |
| $A_{BC}$ | -7.756 | [-8.543, -6.970] |
| $r_{BC}$ | 0.243 | [0.229, 0.257] |
| $A_{ON}$ | -4.641 | [-5.092, -4.191] |
| $r_{ON}$ | 0.189 | [0.177, 0.194] |
| $A_{AB}$ | -9.288 | [-9.675, -8.901] |
| $r_{AB}$ | 0.260 | [0.253, 0.267] |
| $A_{QC}$ | -10.073 | [-10.760, -9.386] |
| $r_{QC}$ | 0.269 | [0.257, 0.282] |

**NOTES.** $A_i$ and $r_i$ are parameter estimates of equation (1), where $i$ denotes the political jurisdiction under consideration. Here, CAN means Canada, BC denotes British Columbia, ON is Ontario, AB is Alberta, and QC is Quebec. Each parameter estimate is provided along with its 95% confidence interval. Each parameter estimate is significant at the 1% level.

**Figure 1.**
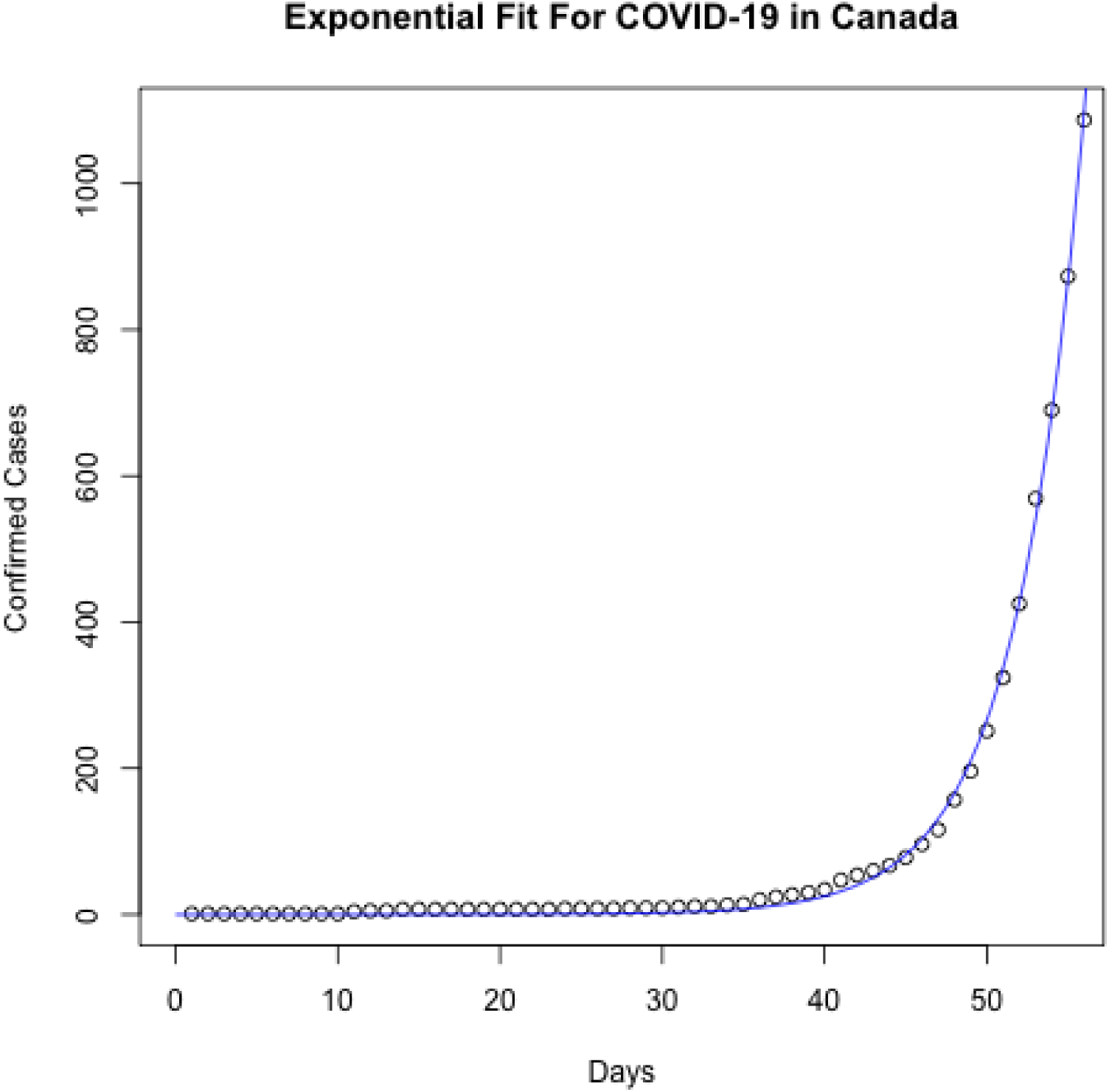
Exponential Fit for Canada.

For policymakers to get a sense of how quickly the number of cases can grow across time, consider Appendix Figure 2. As an illustrative example, consider the following. On March 20, 2020, there were 1,087 cases of SARS-CoV-2 in Canada. One week later, on March 27, 2020, the amount of cases is expected to more than quintuple to over 5,700, based on the exponential model. A week after that, on April 3, 2020, the number of cases is expected to reach 30,131, if current trends do not abate. These numbers will pose significant threats to ICU bed capacity, and the ability of hospitals to provide front-line medical staff with maximum personal protective equipment (PPE).

**Figure 2.**
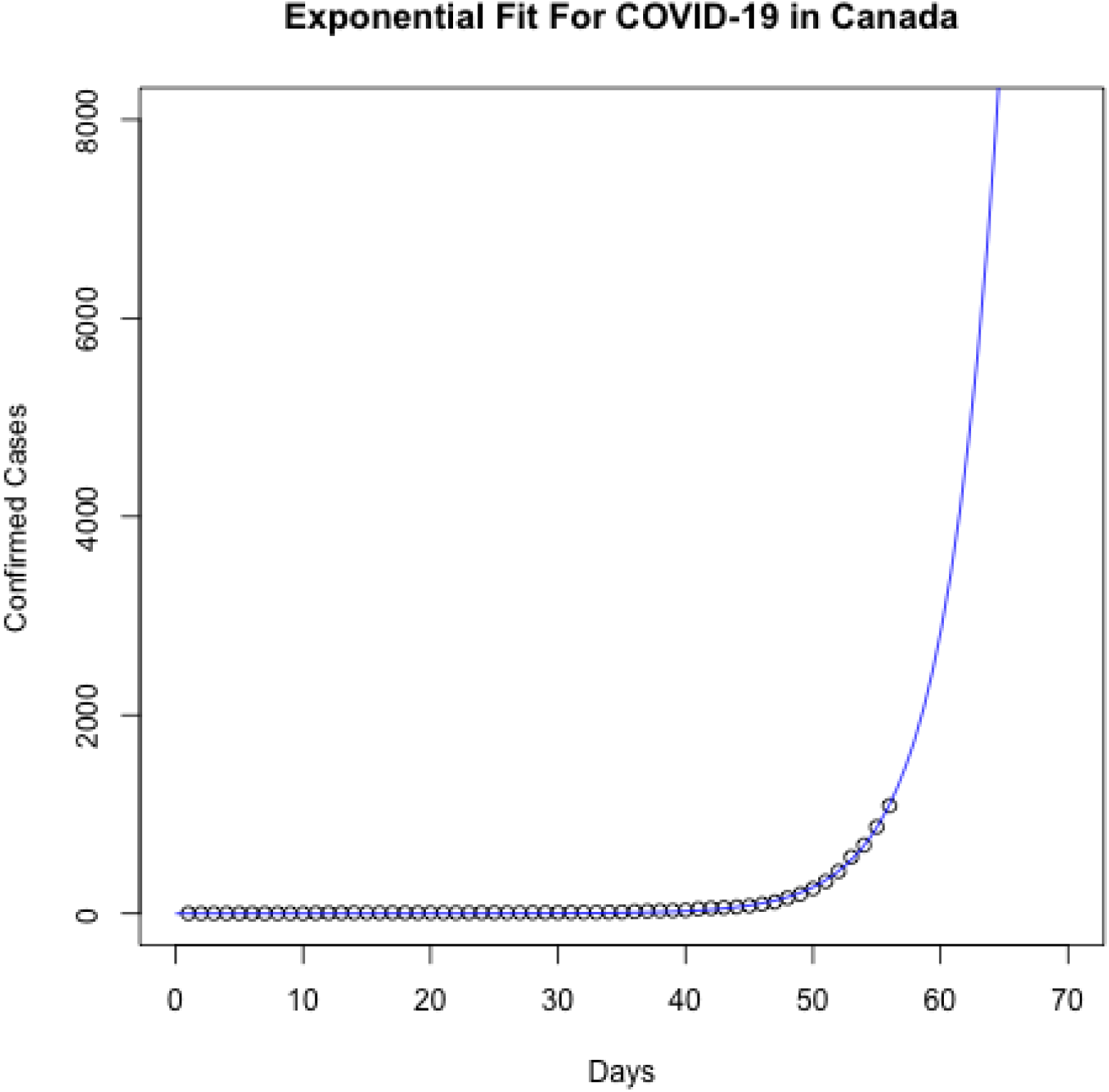
Exponential Fit for Canada.

Canada has ten provinces, four of which seem to be driving this exponential trend: British Columbia, Ontario, Alberta, and Quebec. As of March 20, 2020, 92 percent of Canada’s COVID-19 cases come from these four provinces. These four provinces also contain 86 percent of Canada’s population, and contain a number of other factors that make them more susceptible to disease transmission: densely populated urban areas, international and national travel hubs, and immigration.

The same non-linear least squares method was applied to each individual province’s number of COVID-19 case numbers, with results shown in Appendix Figure 3 (British Columbia), Figure 4 (Ontario), Figure 5 (Alberta), and Figure 6 (Quebec). In each of these provinces, an exponential fit appears to model the data well, strongly suggesting good prediction.

**Figure 3.**
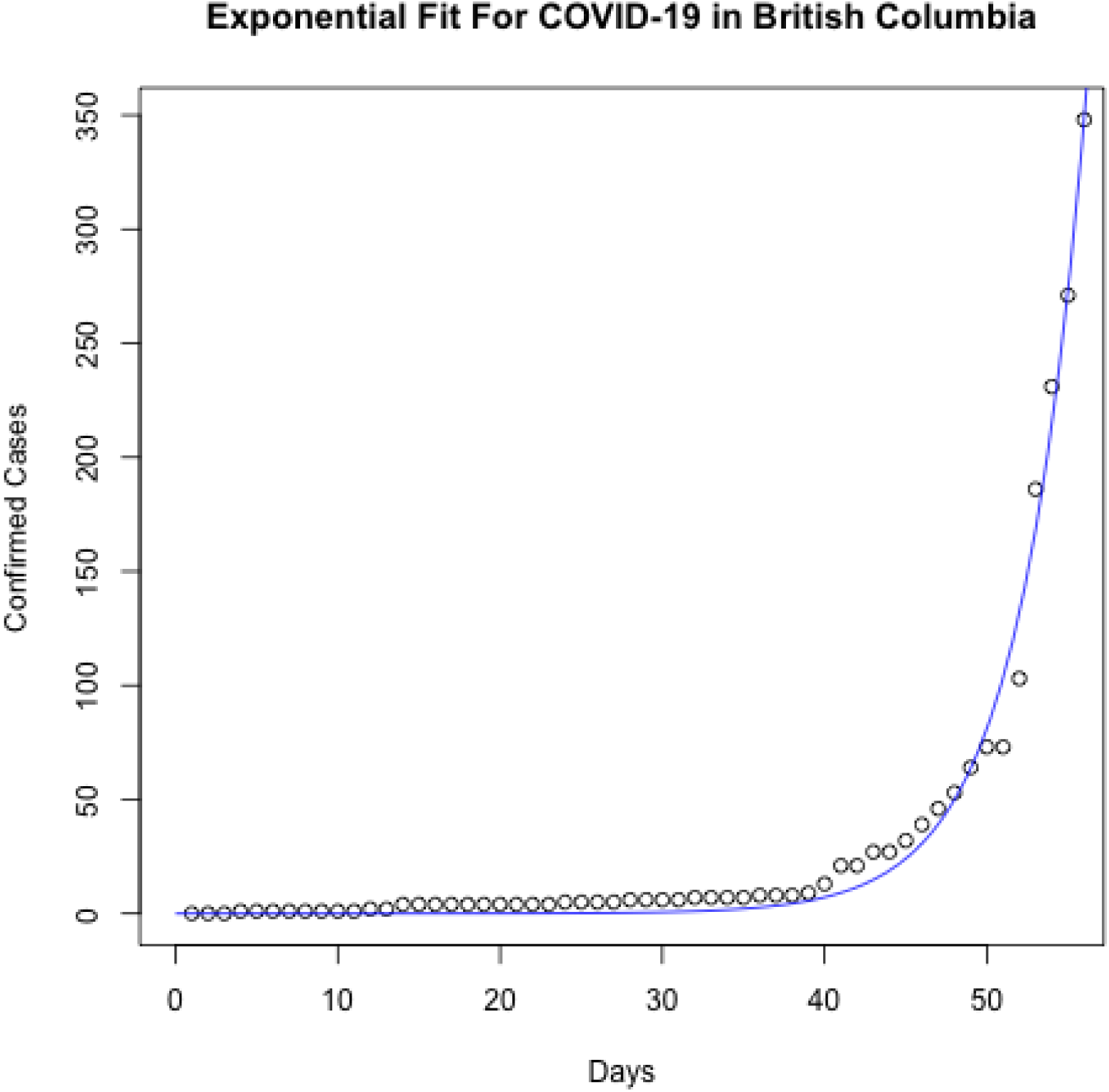
Exponential Fit for Canada.

**Figure 4.**
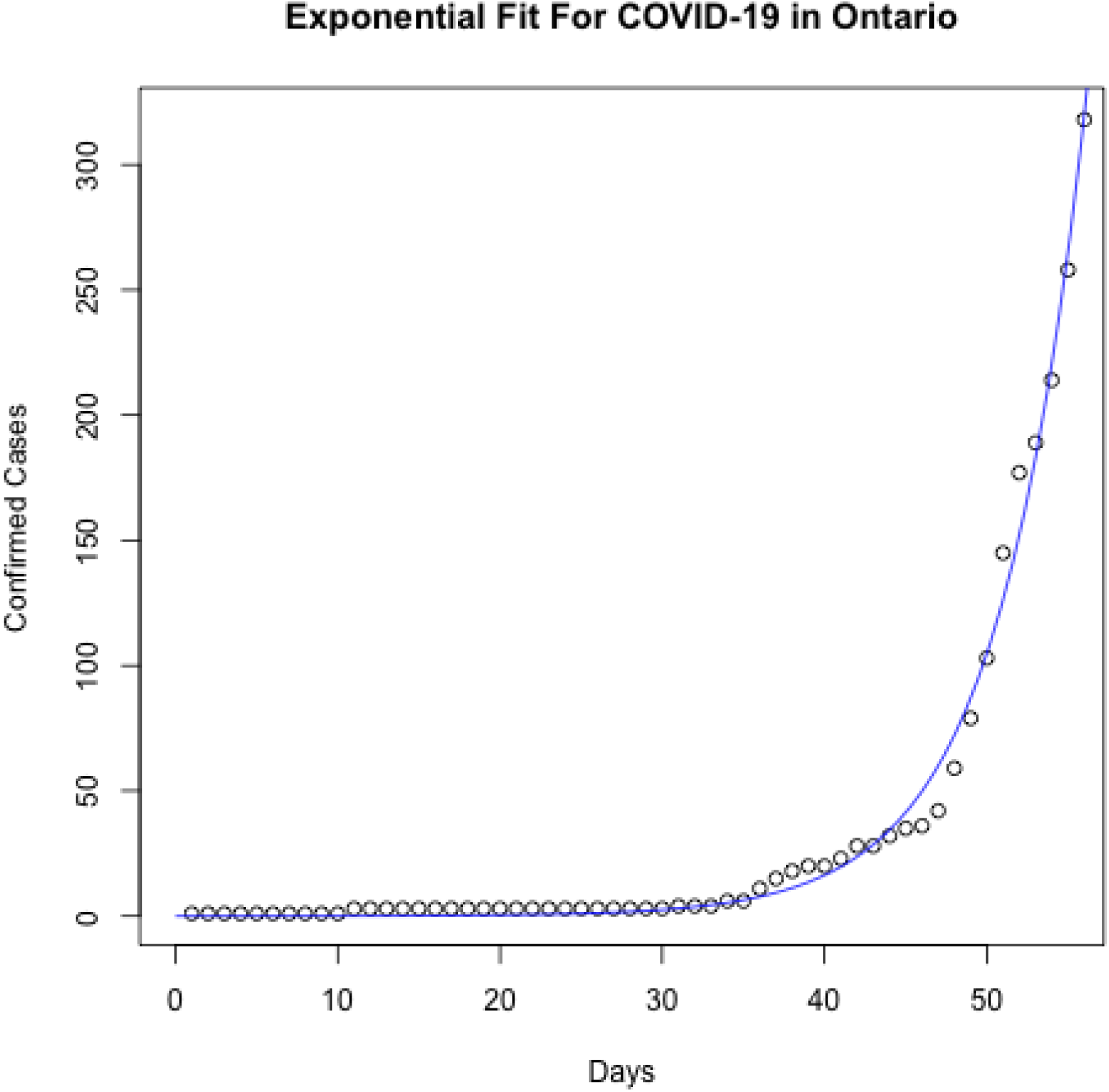
Exponential Fit for Ontario.

**Figure 5.**
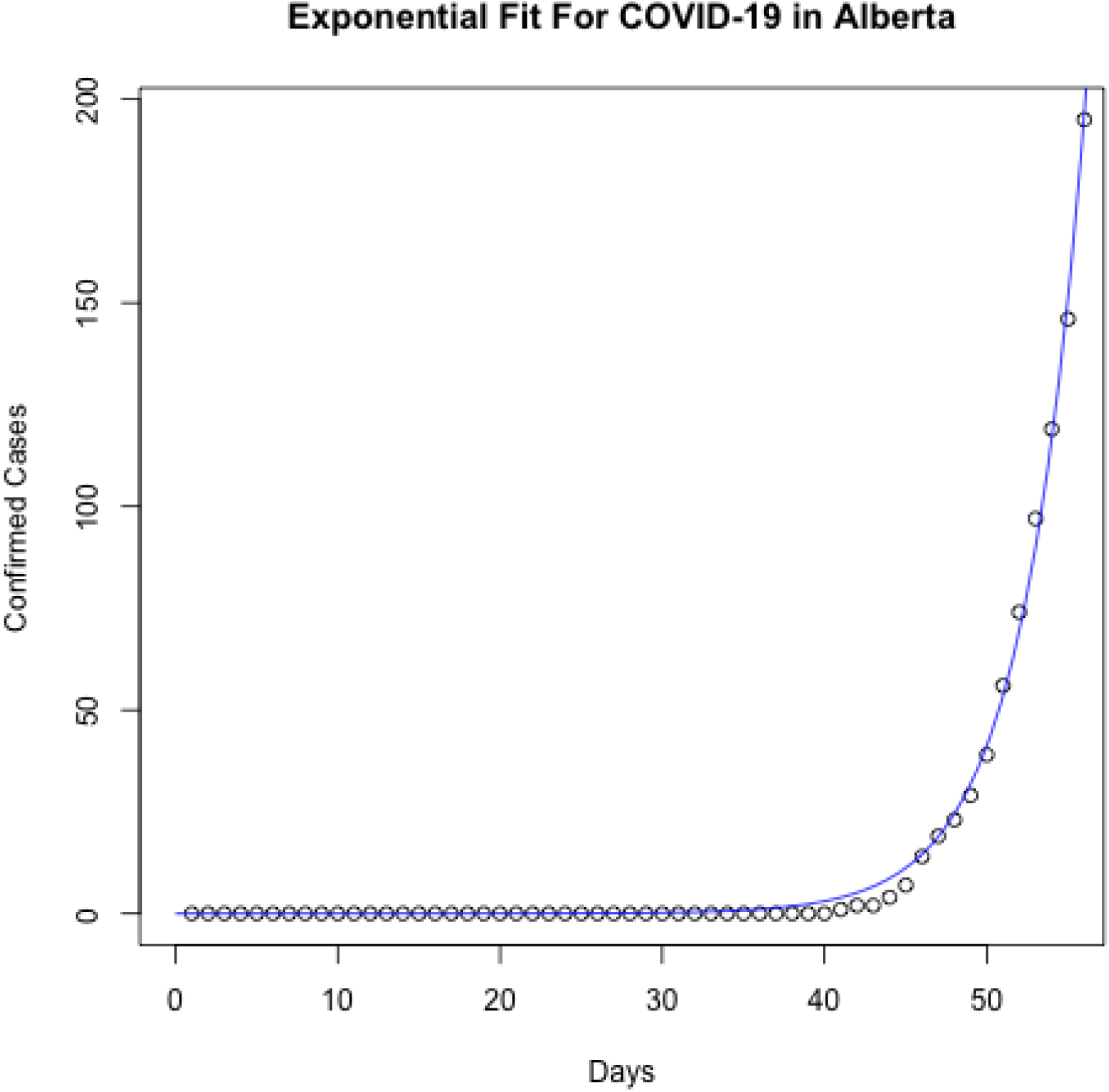
Exponential Fit for Alberta.

**Figure 6.**
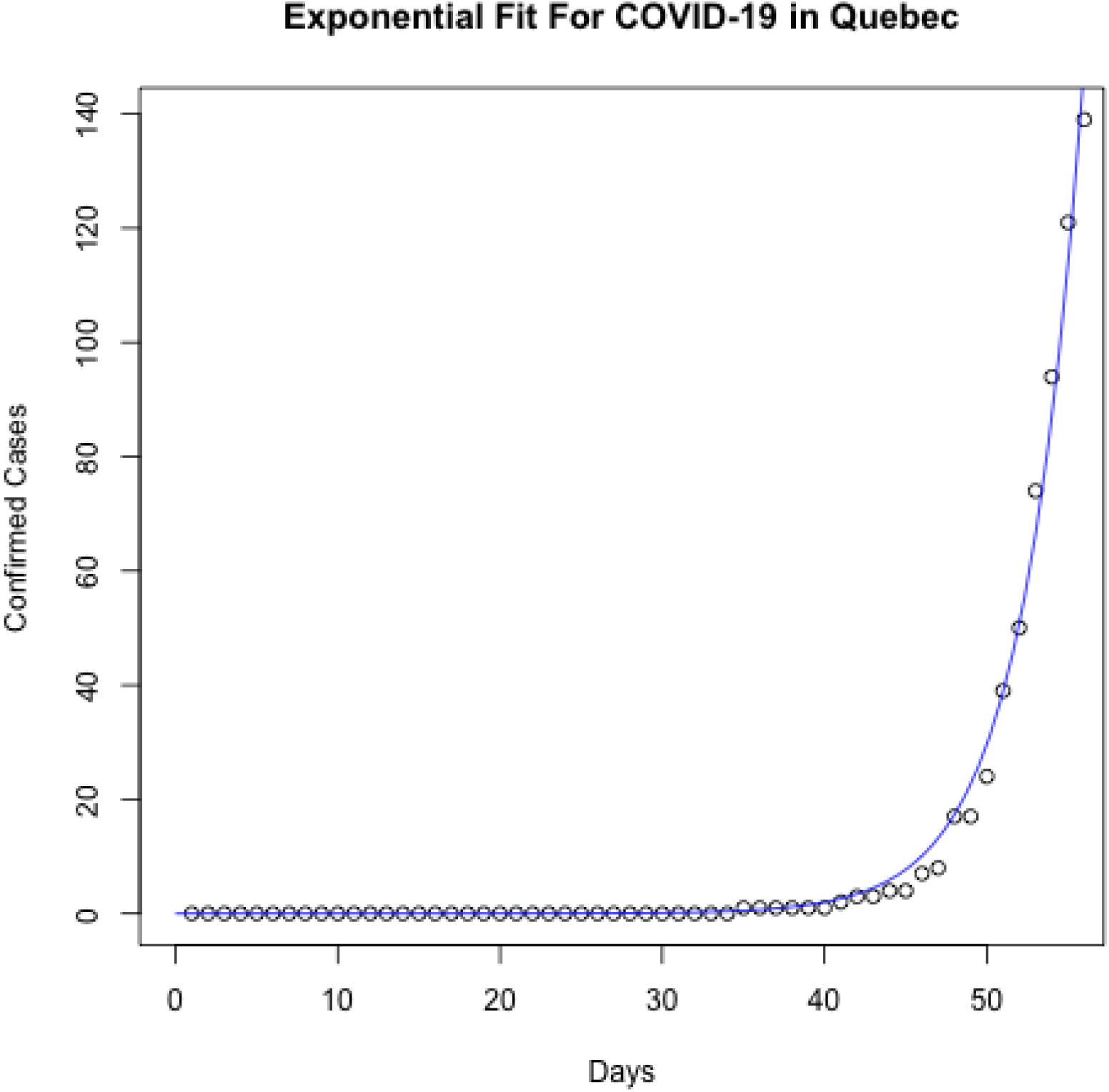
Exponential Fit for Quebec.

## Discussion

As with any discussion of exponential virus growth, it is difficult to predict exactly when the peak of the outbreak will take place in Canada. However our models are consistent with other models of exponential growth, including those from Italy where the exponential growth pattern continues unabated.^5^

Countries in Asia, including Taiwan, Singapore, and Hong Kong, have rapidly brought the COVID-19 outbreak under control, and have important lessons to teach European and North American nations struggling with exponential growth rates. Both these nations have used a multitude of early, aggressive interventions including rapid border controls, social distancing, the innovative and population-wide use of smart phone technology and the ubiquitous wearing of surgical masks in healthcare settings as well as the general population.^10^ Apart from the late and uncertain adoption of social distancing, Canada has not been willing or able to learn from these countries’ experience. In particular, there is not yet any attempt to mobilize smart phone technology or use face masks for all healthcare workers.

Face masks have been shown to reduce infection rates in influenza epidemics and WHO officials have been shown in news media to be wearing face masks whilst addressing news conferences. Smart phone technology adapted to Canadian needs would involve the creation of a national-level app which can be downloaded and used to track COVID-19 cases in real time, provide uniform updates on public policy and announcements; and make simple, consistent messaging available on a national scale.

The ubiquitous use of face masks, at least in healthcare settings is a rapid adoption NPI which can have major impacts on COVID-19 related morbidity and mortality in Canada and elsewhere.^11^

Policymakers in Canada should also implement a rapid roll out of policies that harness the power of smart phone technology^12–15^ and the wearing of face masks by the public. Unfortunately, the messaging in Canada has been counterproductive and has included various versions of “face masks don’t work unless properly used.” This is the same as claiming that hand washing does not work, unless properly carried out.

Both Taiwan and Hong Kong function as forms of democracies, not unlike Canada and the United States. It should therefore be possible to move with deliberate and urgent speed to implement policies that have been shown to work in these Asian countries. Mandating healthcare workers to wear a mask in work places is a good start.

Failure to make bold and rapid moves based on the successful measures that have been implemented in Taiwan and Hong Kong is likely to prove very costly in terms of overloaded ICU resources, public mistrust of health officials and an exponential increase in morbidity and mortality, the latter of which were observed in countries like Italy.

## Data Availability

For code and data, please contact the corresponding author.

## Appendices

## Notes

### Competing Interest Statement

The authors have declared no competing interest.

### Funding Statement

No funding to declare.

